# Impact of policy interventions and social distancing on SARS-CoV-2 transmission in the United States

**DOI:** 10.1101/2020.05.01.20088179

**Authors:** Nickolas Dreher, Zachary Spiera, Fiona M. McAuley, Lindsey Kuohn, John R. Durbin, Naoum Fares Marayati, Muhammad Ali, Adam Y. Li, Theodore C. Hannah, Alex Gometz, JT Kostman, Tanvir F. Choudhri

**Affiliations:** Icahn School of Medicine at Mount Sinai, Department of Neurosurgery; Concussion Management of New York; ProtectedBy.AI

**Keywords:** COVID-19, SARS-CoV-2, Coronavirus, Public Policy, Social Distancing, Non-pharmaceutical Interventions, Stay-at-home Order, Shelter-in-place

## Abstract

**Background:** Policymakers have employed various non-pharmaceutical interventions (NPIs) such as stay-at-home orders and school closures to limit the spread of Coronavirus disease (COVID-19). However, these measures are not without cost, and careful analysis is critical to quantify their impact on disease spread and guide future initiatives. This study aims to measure the impact of NPIs on the effective reproductive number (R_t_) and other COVID-19 outcomes in U.S. states.

**Methods:** In order to standardize the stage of disease spread in each state, this study analyzes the weeks immediately after each state reached 500 cases. The primary outcomes were average R_t_ in the week following 500 cases and doubling time from 500 to 1000 cases. Linear and logistic regressions were performed in R to assess the impact of various NPIs while controlling for population density, GDP, and certain health metrics. This analysis was repeated for deaths with doubling time from 50 to 100 deaths and included several healthcare infrastructure control variables.

**Results:** States that had a stay-at-home order in place at the time of their 500th case are associated with lower average R_t_ the following week compared to states without a stay-at-home order (p < 0.001) and are significantly less likely to have an R_t_>1 (OR 0.07, 95% CI 0.01 to 0.37, p = 0.004). These states also experienced a significantly longer doubling time from 500 to 1000 cases (HR 0.35, 95% CI 0.17 to 0.72, p = 0.004). States in the highest quartile of average time spent at home were also slower to reach 1000 cases than those in the lowest quartile (HR 0.18, 95% CI 0.06 to 0.53, p = 0.002).

**Discussion:** Few studies have analyzed the effect of statewide stay-at-home orders, school closures, and other social distancing measures in the U.S., which has faced the largest COVID-19 case burden. States with stay-at-home orders have a 93% decrease in the odds of having a positive R_t_ at a standardized point in disease burden. States that plan to scale back such measures should carefully monitor transmission metrics.

## Introduction

Severe acute respiratory syndrome coronavirus 2 (SARS-CoV-2) causing coronavirus disease 2019 (COVID-19) was first reported in Wuhan, China in December of 2019.^1^ It quickly spread globally, and was characterized as a pandemic by the World Health Organization (WHO) in March (2020). Local and national governments worldwide have employed a variety of non-pharmaceutical interventions (NPIs) to mitigate the impact of this novel coronavirus. Mandated policies including limitations on mass gatherings, business closures, and stay-at-home orders have aimed to encourage social distancing and flatten the curve.^2–4^

As of April 30, 2020, over 3,249,000 COVID-19 cases have been confirmed worldwide, with more than 1,067,000 cases and 62,000 resulting deaths in the United States.^5^ In an effort to contain the virus, broad shutdowns have resulted in severe economic impacts including 26 million Americans filing for unemployment within a 5 week period.^6^ Simultaneously, there is concern that quarantine puts people at increased risk of domestic violence and severe psychological suffering, as well as physical inactivity, weight gain, behavioral addiction disorders, and insufficient sunlight exposure.^7–12^ It is therefore important to quantify the effects of social distancing measures on disease spread in order to guide future policy decisions which may continue to limit economic security and healthy lifestyles.

Previous modeling studies predicted that ‘social distancing’ policies could be critical in mitigating the spread of COVID-19.^13–19^ Recent reports have begun exploring the effectiveness of social distancing in reducing disease spread at the country level and local county level.^2,20–22^ Mandated NPIs have also been associated with reduced transmission of SARS-CoV-2 in Wuhan.^23^ Further, the United States CDC has demonstrated that social distancing policies have reduced community mobility in Seattle, San Francisco, New York City, and New Orleans.^3^ However, literature exploring the actual effects of various social distancing policies on disease transmission across states in the U.S. remains sparse. Furthermore, efforts to quantify the effects on transmission have not accounted for different stages of disease burden, discounting that the efficacy of policy changes will likely differ if they are instituted in the context of 20 cases or 10,000. This study accounts for the stage of disease spread by selecting a normalized point on the epidemic curve, analyzing each state in the week following its 500th case and assessing how different NPIs influence the burgeoning case load.

## Methods

### Measures

In order to retrospectively analyze metrics of disease spread and mortality, case and death data were compiled up to April 30th, 2020 from the COVID-19 time series made available by *The New York Times*. Daily estimates of the virus’s effective reproduction number (R_t_) were collected for all 50 U.S. states and the District of Columbia from rt.live, which tracks COVID-19 spread and provides state-level estimates of R_t_. Details on the methodology they used to calculate R_t_ are publicly available online.

To standardize the stage of disease spread and minimize the confounding effect of increased caseload on disease transmission across states, these analyses were conducted in the weeks after a state’s 500th case. The 500 case threshold was chosen to ensure that each state had a sustained epidemic while still encompassing almost all states.

The primary outcomes were average R_t_ in the weeks following the 500th case and doubling time from 500 to 1000 cases, both measures of disease transmission. R_t_ is a real-time measure of the basic reproduction number (R_0_), which estimates the number of infections expected from one case interacting with a susceptible population.

A secondary analysis investigated the effects of NPIs on doubling time from 50 to 100 deaths and case fatality rate. Again, the 50 deaths threshold was chosen to ensure that each state had faced enough COVID-19 spread to experience sustained morbidity, while still encompassing most states. Only a minority of states have reached 500 deaths to date, so the threshold used for the case analysis was not applicable. A rough estimate of case fatality rate was calculated by simply dividing deaths by total cases for each state.

In order to better understand effects of NPI on social mobility, and the effects of social mobility on disease spread, social distancing metrics were collected from the COVID-19 community mobility reports made available by Google. These reports compare the average time spent in places of residence based on Google location tracking data compared to the median value, for the corresponding day of the week, during the 5-week period Jan 3–Feb 6, 2020. Averages of these measures were calculated for the week after stay-at-home order to assess the impact of NPI on social distancing. Furthermore, average increase in time spent in residential areas was also calculated for the week before the 500th case to assess the impact of social distancing on disease transmission directly.

### Covariates

We tested the association between five unique policy changes and the change in R_t_: stay-at-home orders, school closures, closure of non-essential businesses, and bans on mass gatherings. Demographic data, including population density, population size, and GDP were obtained from publicly available data for each state and territory and examined as covariates in multivariable models. State-wide health information, including the percentage of state residents with diabetes, chronic obstructive pulmonary disease (COPD), current and ever smokers, and cardiovascular disease, were included to control for potential confounding effect. Lastly, the number of hospital beds and physicians per 1000 people were used to control for state-specific health care capacity. These measures were assessed as covariates in the secondary analysis examining case fatality rate.

### Statistical Analysis

All analyses were complete in R (Version 1.1.442) and Microsoft Excel. Descriptive statistics are reported using means (standard deviation [SD]) and median (interquartile range [IQR]) for normally and non-normally distributed continuous variables, respectively. The Kruskal Wallis test was used to determine differences for non-normally distributed variables. Policy changes were modeled as dichotomous variables distinguishing states that had implemented each order 1) prior to the 500th case in primary analyses and 2) prior to the 50th death in secondary analyses. Univariable linear regression was used to test the association between each policy change and the primary outcome, average R_t_ after a state’s 500th case. Average R_t_ after the 500th case was then dichotomized into values above and below 1 and evaluated in logistic regression. Multivariable models were then built to adjust for demographic, state-wide health, and health care capacity covariates. Kaplan Meier survival analysis and the log-rank sum test were used to identify differences in the time to reach the 1000th case. The average % time spent at home was separated into quartiles and the highest and lowest quartiles were compared. Cox proportional hazards regression was then used to test the association between covariates the risk for reaching 1000 cases. Visual inspection and calculation of the scaled Schonfield residuals were used to confirm the proportional hazard assumption. All analyses were then repeated for case fatality rate and time to 100th death. Multivariable models were built by selecting covariates with p < 0.1 in univariable analyses, backwards eliminating covariates with p > 0.1, and removing collinear variables identified by a variance inflation factor >5.

## Results

As of April 30th, 2020, 48 states and the District of Columbia had reached 500 cases. Of these states, 15 had stay-at-home orders enacted prior to the date of their 500th case (**Table 1**). These locations had a significantly smaller (p = 0.007) median population (1,826,156) compared to states without this policy implemented before reaching 500 cases (5,967,435). There were no statistically significant differences between cohorts in population density, hospital beds per 1000 people, physicians per 1000 people, percent current smokers, percent with COPD, percent with diabetes, or percent with cardiovascular disease.

**Table 1:** Summary of included states and territories (N = 49)

| Variable (Median [IQR]) | All states and District of Columbia<br>N = 49 | States without stay-at-home order at 500 cases<br>N = 34 | States with stay-at-home order at 500 cases<br>N = 15 | p |
| --- | --- | --- | --- | --- |
| Population | 4,645,184<br>[1,952,570 to 7,797,095] | **5,967,435**<br>**[3,149,705 to 9,767,915]** | **1,826,156**<br>**[1,358,518 to 4,400,391]** | **0.007\*** |
| Population density | 112.82<br>[56.93 to 228.02] | 110.44<br>[57.11 to 225.63] | 113.96<br>[56.48 to 253.72] | 0.8 |
| Hospital beds per 1000 people | 2.50<br>[2.10 to 3.10] | 2.55<br>[2.10 to 3.18] | 2.10<br>[1.95 to 2.60] | 0.09 |
| Physicians per 1000 people | 2.74<br>[2.41 to 3.14] | 2.55<br>[2.10 to 3.18] | 3.08<br>[2.62 to 3.29] | 0.1 |
| % Current smokers | 17.00<br>[14.60 to 19.30] | 17.15<br>[14.80 to 19.18] | 16.10<br>[14.70 to 19.30] | 0.9 |
| % COPD | 6.70<br>[5.60 to 8.30] | 6.50<br>[5.38 to 8.28] | 6.90<br>[5.95 to 8.30] | 0.7 |
| % Diabetes | 11.00<br>[9.90 to 12.50] | 10.90<br>[9.75 to 12.57] | 11.00<br>[10.25 to 12.35] | 0.8 |
| % Cardiovascular Disease | 4.30<br>[3.70 to 5.00] | 4.30<br>[3.80 to 5.07] | 3.90<br>[3.65 to 5.00] | 0.8 |

Abbreviations: IQR = interquartile range, COPD = chronic obstructive pulmonary disease

### NPI effects on disease spread

48 states and the District of Columbia were included in this analysis. Alaska and Montana were excluded because they had not yet reached 500 confirmed COVID-19 cases as of April 30th, 2020. Average R_t_ for all included territories the week prior to implementing stay-at-home orders (R_t_ = 1.256) compared to the week following (R_t_ = 1.088) was reduced -13.3% (absolute change = -0.1673, SD = 0.070).

States with stay-at-home orders preceding the date of their 500th case were negatively associated with average R_t_. (ß = -0.15, 95% CI -0.23 to -0.07, p < 0.001, **Table 2**). Educational facilities closure (ß = -0.17, 95% CI -0.30 to -0.05, p = 0.009), non-essential business closure (ß = 0-.13, 95% CI -0.30 to -0.05, p = 0.002), and average % time spent at home the week before (ß = -0.02, 95% CI -0.02 to -0.01, p < 0.001) were also associated with a significant reduction in R_t_ compared to states without these policies the week following 500 cases.

From days 8 to 14 after the 500th case date, implementation of stay-at-home order (ß = -0.09, 95% CI -0.15 to -0.04, p < 0.002), educational facilities closure (ß = -0.12, 95% CI -0.21 to -0.04, p = 0.006), non-essential business closure (ß = -0.05, 95% CI -0.13 to 0.03, p = 0.004), and average % time spent at home the week before (ß = -0.01, 95% CI -0.01 to 0.00, p = 0.005) implemented prior to the 500th case date were associated with a significant reduction in R_t_ compared to controls.

In multivariable analyses, average percent time spent at home during the week before remained a significant predictor of reduction in R_t_ (ß = -0.01, 95% CI -0.02 to -0.01, p=0.001) when adjusting for stay-at-home orders. However, when evaluating the R_t_ with a one week delay after the 500th case, average percent time spent at home was no longer associated (ß = -0.01, 95% CI -0.01 to -0.00, p=0.07). Other covariates, including school closures, limitations on mass gatherings, non-essential business closure, population density, and population size were not found to be associated with R_t_ when evaluated alongside average time spent at home and therefore were not included in the multivariable model.

We then dichotomized R_t_ into values above and below 1 and repeated the analysis with a univariable logistic regression model. In this analysis, implementing a stay-at-home order was associated with a 93% decrease in the odds of having a positive R_t_ in the week immediately following the 500th case (OR 0.07, 95% CI 0.01 to 0.37, p=0.004). The following week also experienced an 84% decrease in the odds of having an average R_t_ greater than 1 (OR 0.16, 95% CI 0.04 to 0.58, p=0.008).

**Table 2:** Linear and logistic regressions assessing the impact of non-pharmaceutical interventions on R_t_ following 500 cases

| Covariate | $\beta$ (95% CI) | p | OR (95% CI) | p |
| --- | --- | --- | --- | --- |
| Week immediately following 500th Case (days +1 to +7) |  |  |  |  |
| Stay-at-home order | **-0.15 (-0.23 to -0.07)** | **<0.001\*** | **0.07 (0.01 to 0.37)** | **0.004\*** |
| Limitation on mass gatherings | -0.08 (-0.20 to 0.04) | 0.2 | Limited sample size |  |
| Educational facilities closure | **-0.17 (-0.30 to -0.05)** | **0.009\*** | Limited sample size |  |
| Non-essential business closure | **-0.13 (-0.21 to -0.05)** | **0.002\*** | **0.09 (0.01 to 0.43)** | **0.006\*** |
| Average % time spent at home in the week before | **-0.02 (-0.02 to -0.01)** | **<0.001\*** | 0.82 (0.64 to 0.99) | 0.07 |
| One-week delay from 500th case (days +8 to +14) |  |  |  |  |
| Stay-at-home order | **-0.09 (-0.15 to -0.04)** | **0.002\*** | **0.16 (0.04 to 0.58)** | **0.008\*** |
| Limitation on mass gatherings | -0.05 (-0.13 to 0.03) | 0.2 | 0.18 (0.01 to 1.15) | 0.1 |
| Educational facilities closure | **-0.12 (-0.21 to -0.04)** | **0.006\*** | Limited sample size |  |
| Non-essential business closure | **-0.05 (-0.13 to 0.03)** | **0.004\*** | **0.21 (0.05 to 0.72)** | **0.02\*** |
| Average % time spent at home in the week before | **-0.01 (-0.01 to -0.00)** | **0.005\*** | **0.82 (0.67 to 0.95)** | **0.02\*** |

**Figure 1:**
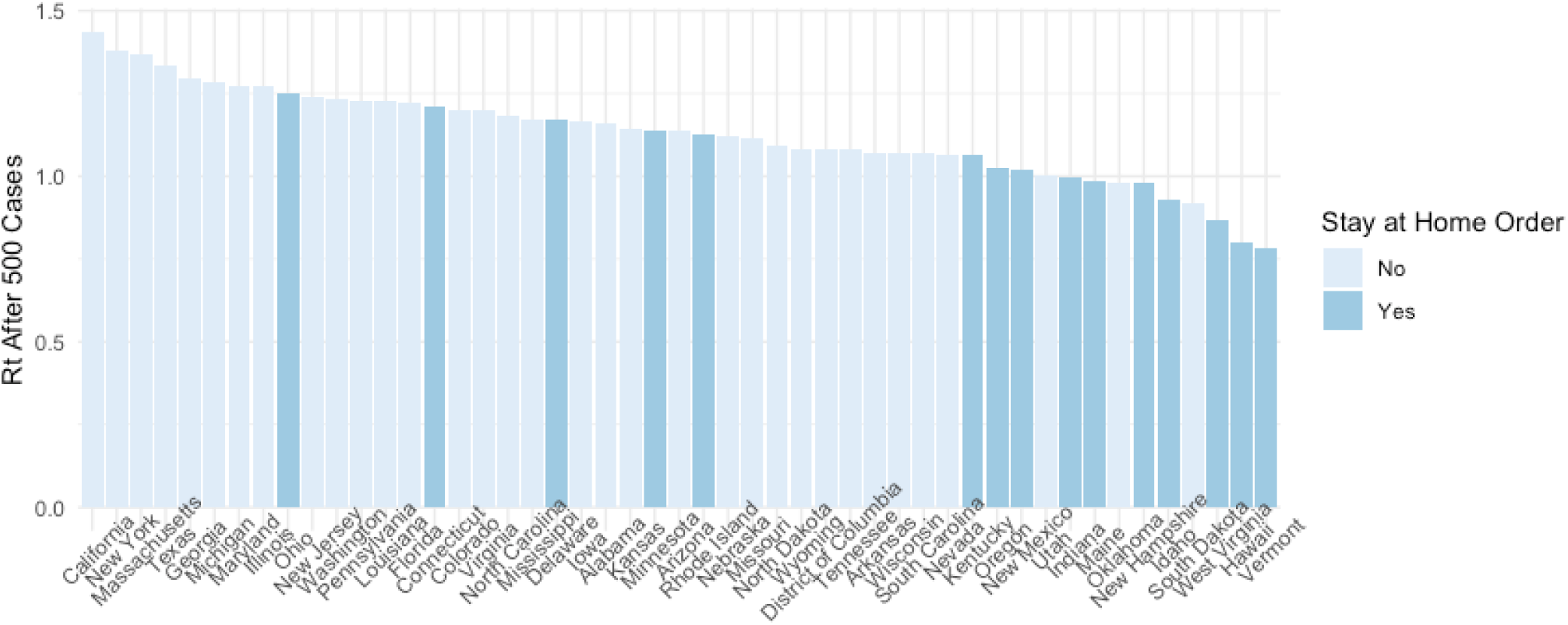
Average R_t_ during the week following the 500th case by each state.

In Kaplan Meier analyses, implementation of a stay-at-home order prior to the date of 500 cases was associated with a decreased probability of reaching 1000 cases within 5 days (log rank sum, p = 0.02). Similarly, in cox proportional hazards regression, stay-at-home orders correlated with an increase in time to reach 1000 cases (OR = 0.35, CI 0.17 to 0.92, p = 0.004, **Table 3, Figure 2**). States in the highest quartile of average percent time spent at home were also less likely to reach 1000 cases (log rank sum, p<0.001, HR 0.18, 95% CI 0.06 to 0.53, p=0.002). Other distancing measures did not affect the time from 500 to 1000 cases.

**Figure 2:**
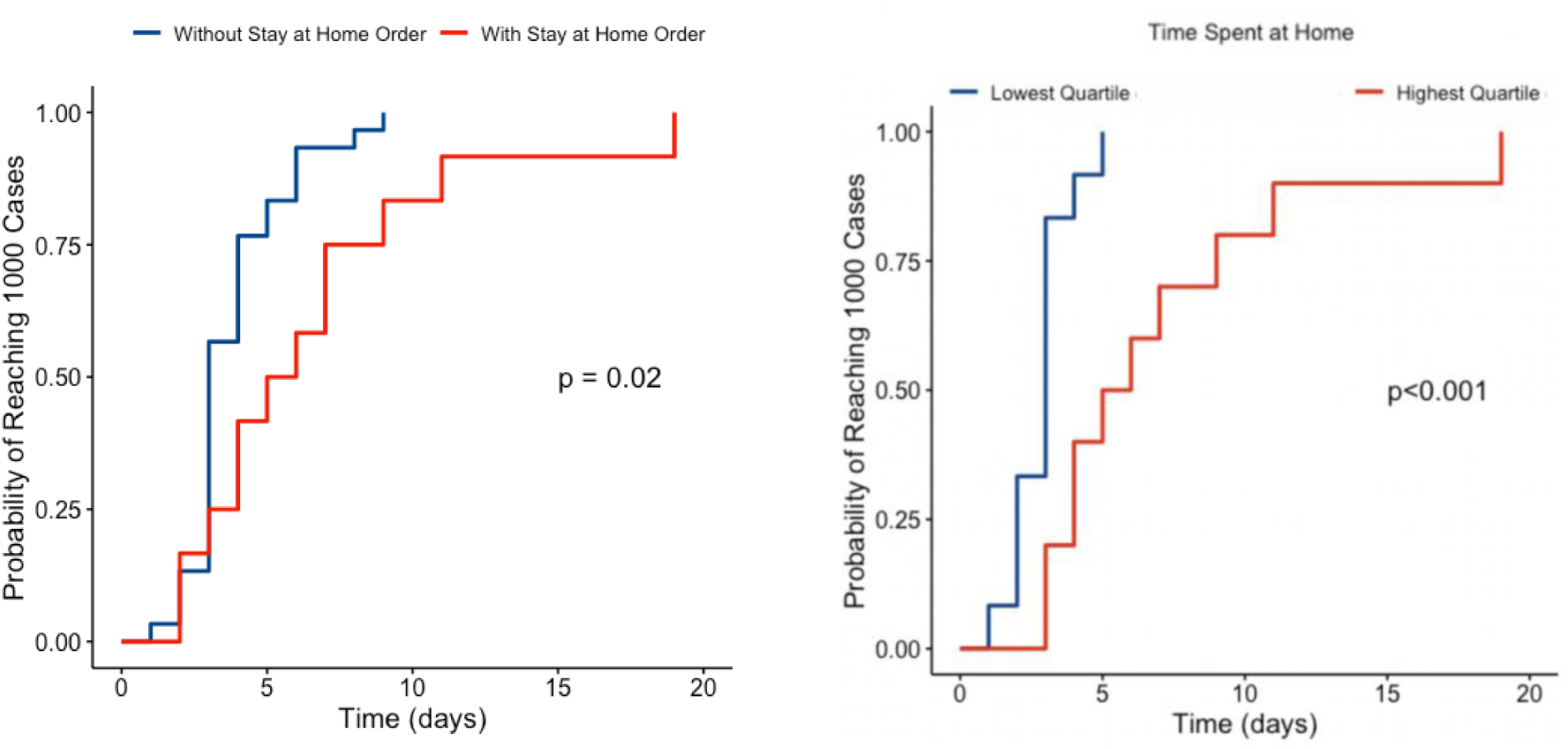
Hazards curve demonstrating the probability of reaching 1000 cases separated by (A) states with and without a stay-at-home order prior to the 500th case and (B) the highest vs. lowest quartile of % time spent at home based on Google mobility data.

**Table 3:** Cox proportional hazards regression for time to event analysis

|  | Time to 1000th Case |  |
| --- | --- | --- |
| Covariate | Hazard ratio (95% CI) | p |
| Stay-at-home order | **0.35 (0.17 to 0.72)** | **0.004\*** |
| Educational facilities closure | 0.63 (0.25 to 1.63) | 0.3 |
| Non-essential business closure | 0.55 (0.28 to 1.10) | 0.08 |
| Limitation on mass gatherings | 0.75 (0.31 to 1.79) | 0.5 |
| Average % time spent at home (Q4 vs. Q1) | **0.18 (0.06 to 0.53)** | **0.002\*** |

### NPI effects on deaths

In linear regression, this study found that none of the included policies (stay-at-home orders, school closures, bans on mass gatherings, or closure of non-essential businesses) were associated with a decrease in case fatality rate (CFR). In Kaplan Meier event analysis, stay-at-home orders were non-significant in predicting time from 50 deaths to 100 deaths (**Figure 3**).

**Figure 3:**
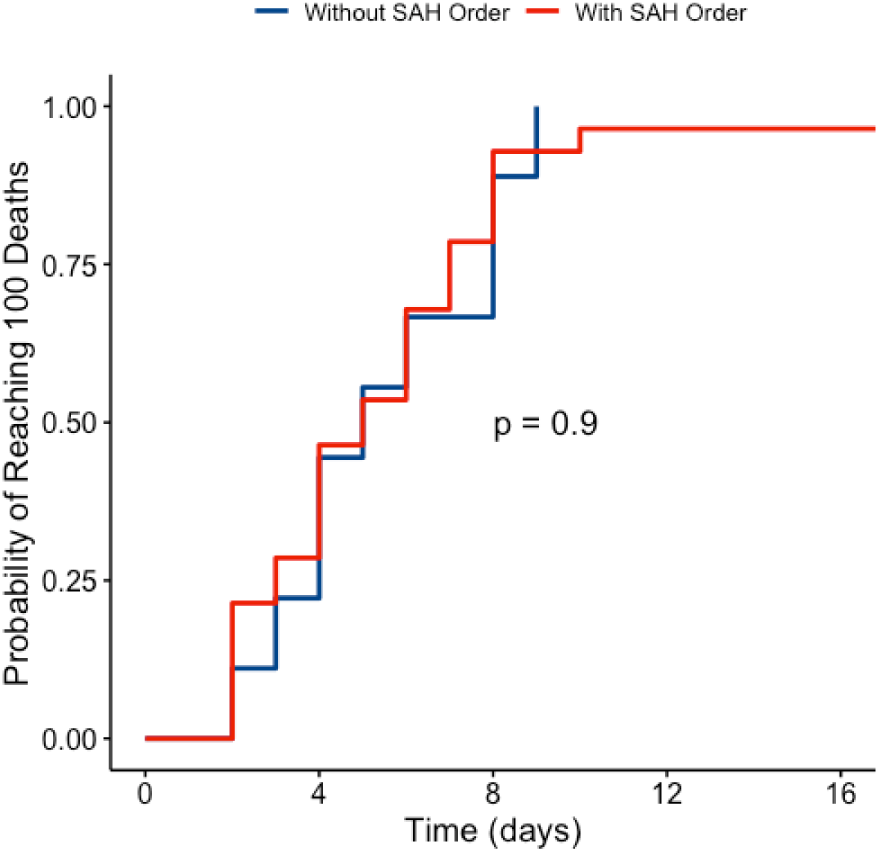
Hazard curve showing the probability of reaching 100 deaths separated by states with and without a stay-at-home order prior to the 50th death

### NPI Interaction with Social Distancing

After the implementation of state-wide stay-at-home orders, the average amount of time spent at home increased by 29.2% relative to the week prior to the order. This translates to an average absolute increase of 4.18% in time spent at home in the week following a stay-at-home order when compared to the previous week. School closures, non-essential business closures, and limitations on mass gatherings led to absolute increases of 10.2%, 5.3%, and 8.1%, respectively.

**Figure 4:**
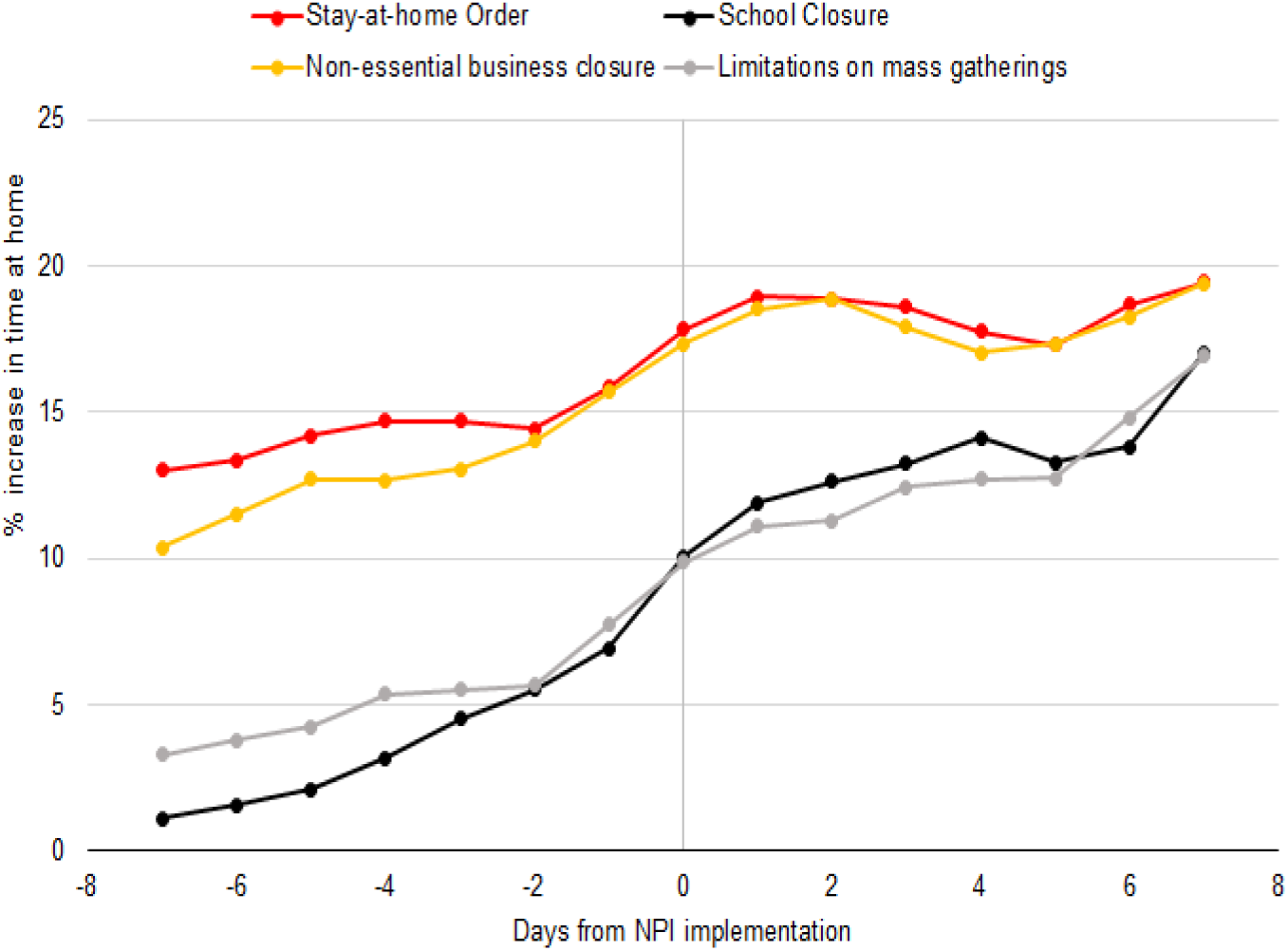
Time spent in residential areas before and after stay-at-home order

## Discussion

This study analyzes state-level transmission rates of COVID-19 after each state’s 500th case, grouping them according to policies implemented prior to their date of 500 cases, in order to determine the efficacy of various social distancing measures. In states that implemented a stay-at-home order prior to reaching 500 cases, we observe a significant decrease in the effective viral transmission rate. Subsequent multivariable analyses indicate that this effect may have been driven by a state-wide increase in the amount of time spent at home. We also determine that enacting stay-at-home orders prior to the 500th case significantly increased the time it took to reach 1000 cases but find no association between social distancing policies and deaths or case fatality rate.

### Context and contribution

Early data to support the efficacy of various NPI measures in reducing SARS-CoV-2 spread has relied mostly on model-based analysis rather than empirical data derived from observations in the real world.^13–19^ As cases have accumulated around the world, it has become increasingly possible to retrospectively assess the impact of NPIs on measured outcomes, as presented in this study. Previous characterization of disease burden across research studies and news sources has largely relied on the metrics of cumulative case and death counts; however, these metrics are unidirectional and do not account for bidirectional changes in the rate of viral transmission over time, a much more powerful metric for predicting an epidemic’s trajectory. In this study, we examine effective reproduction number (R_t_) as the primary metric of disease burden, which describes the virus’s transmission potential in real-time and can thus account for the impact of contextual changes in policy and behavior on disease spread.

Retrospective analyses of NPI impact on disease spread, to date, have primarily consisted of cross-country analyses or focused on outcomes in China.^2,20,21,23–25^ In one comparison of 20 countries, Banholzer et al. found public venue closures to be the most effective NPI in reducing new cases, followed by public gathering bans, non-essential business closures, and international travel restrictions, with school closures decreasing case count minimally. Interestingly, they found ‘lockdowns’ to be among the least effective policies in mitigating disease spread.^2^ Analysis at the city level in China has also associated comprehensive social distancing measures with preventing disease spread.^23,24^ In Wuhan, a reduction in R_t_ was shown to chronologically follow implementation of traffic restrictions, home confinement, centralized quarantine, and other social distancing measures.^23^

The United States presents unique challenges in epidemiological management due to its governmental emphasis on state and local autonomy. As such, an analysis of the pandemic’s impact in the U.S. should account for potentially different trajectories across states and at the local level. Outside of China, local-level studies exploring the effectiveness of NPIs have been scarce, and more granular analysis across U.S. states is currently warranted.^22, 26–27^ Ebell & Bagwell-Adams compared differences in social distancing measures employed by counties in the state of Georgia. They demonstrated that Clarke County, which implemented a shelter in place policy two weeks before it was adopted at the state level, had increased case doubling time compared to surrounding counties and the state as a whole.^22^ Siedner et al. performed a time-series analysis to compare disease spread before and after statewide social distancing policies were put in place, and found that decreases in epidemic growth rates were shown to occur four days after implementation of each state’s first social distancing policy.^28^ However, in this section of their analysis the authors did not differentiate between alternate social distancing measures. Additionally, once an initial policy was in place, they found no significant effect of further enacting statewide lockdowns.^28^ In this study, we compare the effects of different policies, finding stay-at-home orders to be most effective in reducing transmission. Furthermore, by normalizing disease burden across states to 500 cases, we standardize the time point in each state’s outbreak.

Lasry et al. used cell phone data from SafeGraph to assess the relationship between various social distancing policies and percentage of mobile devices leaving home in four major U.S. cities.^3^ They found that combinations of multiple social distancing policies, including limits on gatherings and school closures, significantly reduced mobility. Stay-at-home orders further reduced movement in their study as well.^3^ By including cell phone tracking data made publicly available by Google, this study directly assesses the connection between mobility and virus transmission at the state level. In agreement with Lasry et al., we demonstrate that stay-at-home orders significantly increase the amount of time people spend at home.^3^ Further, our multivariable linear regression analysis, which demonstrates that percent time spent at home was the most significant modulator of R_t_, indicates that the primary driving factor in reducing viral transmission was limiting mobility. In conjunction, these results provide evidence that NPIs can be useful in controlling COVID-19 outbreak by effectively reducing social mobility.

### Differing Effects of NPIs

In our analysis, we found that stay-at-home order, the strictest policy included in our models, had the most significant effect on disease spread. This measure both reduced transmission rate and increased doubling time from 500 to 1000 cases within states. Comparatively, mass gathering restrictions had the least effect on reducing R_t_ across states. As several states across the U.S. prepare to ease social distancing restrictions in the coming weeks, our results suggest that mass gathering restrictions alone may have less of an effect in maintaining R_t_ values below 1. Careful monitoring of R_t_ values in these states may be necessary to proactively identify and control potential recurrent outbreaks.

In order to assess the efficacy of stay-at-home orders at different points in disease outbreak, we also compared states by number of confirmed COVID-19 cases at the time this policy went into effect. We found that reduction in average R_t_ the week following stay-at-home order was consistent across variation in number of cases at the time of policy implementation. States benefited from similar reduction in R_t_ regardless of how many confirmed cases they had before their stay-at-home orders went into effect. However, this finding does not imply that timing of stay-at-home order is unimportant, since high R_t_ in the weeks prior will contribute to greater overall caseload. Furthermore, when looking at more recent R_t_ averages for the week of April 23rd to April 30th, states that have yet to implement a state-wide stay-at-home order currently have amongst the highest values in the country, accounting for four of the eight states with an average R_t_ > 1, suggesting that they have not yet successfully contained the virus.

Our analysis found no significant correlation between mobility or social distancing policy and time from 50 to 100 deaths. This lack of association may be a result of studying outcomes early on in each state’s disease outbreak. During this relatively early timeframe, states may not have reached hospital capacity yet. Future studies that look at death rates later on may find that social distancing measures help prevent overflow of healthcare systems, and therefore reduce fatality. At this time, more longitudinal data is warranted to more accurately characterize the relationship between social distancing efforts and these lagging indicators of disease burden.

### Limitations

Our study has a number of important limitations to consider. First, our state-level analysis may miss variation at the county level. Individual counties may have implemented social distancing measures before mandated state-wide, thus states considered to lack certain policies at the time of 500 cases may in reality have been benefiting from more localized control. Similarly, county variation in COVID-19 cases, resulting deaths, population density, and other demographic factors were not accounted for. Future analyses should consider county-level data to account for these local variations.

Our mobility results are further limited by potential flaws in Google’s publicly available phone data that this study relies on for mobility analyses. As noted by Lasry et al., data that tracks phones, not people, are subject to distortion by individuals with multiple devices and people leaving home without their phones.^3^ Further, these data do not differentiate between individuals leaving home but remaining distanced from others and people who ignore social distancing guidelines altogether while in public. Finally, our analysis focused exclusively on social distancing policies, and did not account for other transmission preventing NPI that states may have employed such as requiring masks.

Lastly, though rates of testing have been noted to vary widely between states and serve as a potentially confounding variable, the model used to calculate R_t_ values analyzed here corrects for these state-wide differences in testing. The R_t_ model also accounts for variation in serial interval and delay between symptom onset and a positive test result; however, it does not account for any period in which individuals are infectious but asymptomatic, which mounting evidence suggests is an important factor in SARS-CoV-2 dynamics. As such, future analyses of R_t_ should be calibrated with this in mind.

## Conclusions

Reducing COVID-19 spread to alleviate overburdened healthcare systems has become an international priority and understanding the effectiveness of policy interventions is paramount. Disease modeling has indicated that social distancing is a critical measure to achieve this goal, but few studies have validated this finding with emerging case data. Furthermore, few have analyzed epidemiology across states in the country with the largest disease burden, the United States. This study indicates that stay-at-home orders, limitations on mass gatherings, educational facility closures, and non-essential businesses closures are all effective measures at reducing transmission rates thereby *flattening the curve*. Among these policies, stay-at-home orders had the largest effect, and as states aim to step down from such policies metrics of disease transmission should be carefully monitored to limit recurrent outbreaks. Ultimately, adherence to social distancing appears to be the driving force behind these policies, as states with stay-at-home orders but poor adherence were found to experience similar outcomes to those without such policies. By more rigorously characterizing the state-level strategies that have proved most effective at reducing disease burden, this study aims to provide stakeholders with a more standardized, data-driven framework to guide future policy decisions.

## Data Availability

Data was obtained from publicly available data sources mentioned in our manuscript.

## Notes

### Competing Interest Statement

The authors have declared no competing interest.

### Funding Statement

No external funding was received.

## References

1. Li Q, Guan X, Wu P, et al. Early Transmission Dynamics in Wuhan, China, of Novel Coronavirus-Infected Pneumonia. N Engl J Med 2020; 382(13): 1199–207.

2. Banholzer N, van Weenen E, Kratzwald B, et al. Estimating the impact of non-pharmaceutical interventions on documented infections with COVID-19: A cross-country analysis. medRxiv 2020.

3. Lasry A, Kidder D, Hast M, et al. Timing of community mitigation and changes in reported COVID-19 and community mobility—four US metropolitan areas, February 26–April 1, 2020: Centers for Disease Control and Prevention, 2020.

4. Institute for Health Metrics and Evaluation. COVID-19 resources. 2020, April 30. http://www.healthdata.org/covid (accessed April 30 2020).

5. Johns Hopkins University & Medicine. COVID-19 Dashboard by the Center for Systems Science and Engineering (CSSE) at Johns Hopkins University (JHU). 2020, April 30. https://coronavirus.jhu.edu/map.html (accessed April 30 2020).

6. Reinicke C. Here’s what 5 economists are saying about unemployment after 26 million Americans filed jobless claims in just 5 weeks. Business Insider. 2020, April 23.

7. Taub A. A New Covid-19 Crisis: Domestic Abuse Rises Worldwide. The New York Times. 2020, April 6.

8. Brooks SK, Webster RK, Smith LE, et al. The psychological impact of quarantine and how to reduce it: rapid review of the evidence. The Lancet 2020.

9. Chaturvedi SK. Covid-19, Coronavirus and Mental Health Rehabilitation at Times of Crisis. Journal of Psychosocial Rehabilitation and Mental Health 2020: 1.

10. Chatterjee SS, Malathesh BC, Mukherjee A. Impact of COVID-19 pandemic on pre-existing mental health problems. Asian Journal of Psychiatry 2020.

11. Banerjee D. The COVID-19 outbreak: Crucial role the psychiatrists can play. Asian journal of psychiatry 2020; 50: 102014.

12. Lippi G, Henry BM, Bovo C, Sanchis-Gomar F. Health risks and potential remedies during prolonged lockdowns for coronavirus disease 2019 (COVID-19). Diagnosis 2020; 1 (ahead-of-print).

13. Prem K, Liu Y, Russell TW, et al. The effect of control strategies to reduce social mixing on outcomes of the COVID-19 epidemic in Wuhan, China: a modelling study. The Lancet Public Health 2020.

14. Ferguson N, Laydon D, Nedjati Gilani G, et al. Report 9: Impact of non-pharmaceutical interventions (NPIs) to reduce COVID19 mortality and healthcare demand. London, UK: Imperial College, 2020.

15. Paternina-Caicedo AJ, Choisy M, Garcia-Calavaro C, et al. Social interventions can lower COVID-19 deaths in middle-income countries. medRxiv 2020.

16. Cano OB, Morales SC, Bendtsen C. COVID-19 Modelling: the Effects of Social Distancing. medRxiv 2020.

17. Kissler SM, Tedijanto C, Lipsitch M, Grad Y. Social distancing strategies for curbing the COVID-19 epidemic. medRxiv 2020.

18. Matrajt L, Leung T. Evaluating the effectiveness of social distancing interventions against COVID-19. medRxiv 2020.

19. Koo JR, Cook AR, Park M, et al. Interventions to mitigate early spread of SARS-CoV-2 in Singapore: a modelling study. The Lancet Infectious Diseases 2020.

20. McGrail DJ, Dai J, McAndrews KM, Kalluri R. Enacting national social distancing policies corresponds with dramatic reduction in COVID19 infection rates. medRxiv 2020: 2020.04.23.20077271.

21. Chen Y, Feng Y, Yan C, Zhang X, Gao C. Modeling COVID-19 Growing Trends to Reveal the Differences in the Effectiveness of Non-Pharmaceutical Interventions among Countries in the World. medRxiv 2020.

22. Ebell M, Bagwell-Adams G. Mandatory social distancing associated with increased doubling time: an example using hyperlocal data. medRxiv 2020.

23. Pan A, Liu L, Wang C, et al. Association of public health interventions with the epidemiology of the COVID-19 outbreak in Wuhan, China. Jama 2020.

24. Ruan L, Wen M, Zeng Q, et al. New measures for COVID-19 response: a lesson from the Wenzhou experience. Clinical Infectious Diseases 2020.

25. Sanche S, Lin Y, Xu C, Romero-Severson E, Hengartner N, Ke R. High Contagiousness and Rapid Spread of Severe Acute Respiratory Syndrome Coronavirus 2. Emerging infectious diseases 2020; 26(7).

26. Rotondi V, Andriano L, Dowd JB, Mills MC. Early evidence that social distancing and public health interventions flatten the COVID-19 curve in Italy. OSF Preprints 2020; 26.

27. Cowling BJ, Ali ST, Ng TW, et al. Impact assessment of non-pharmaceutical interventions against coronavirus disease 2019 and influenza in Hong Kong: an observational study. The Lancet Public Health 2020.

28. Mark J Siedner, Guy Harling, Zahra Reynolds, Rebecca F Gilbert, Atheendar Venkataramani, Alexander C Tsai. Social distancing to slow the U.S. COVID-19 epidemic: an interrupted time-series analysis. medRxiv 2020: 2020.04.03.20052373

